# The impact of COVID-19 pandemic on the provision of ambulatory care for patients with chronic neurological diseases in Japan: evaluation of an administrative claims database

**DOI:** 10.1101/2021.05.10.21256951

**Authors:** Kenichiro Sato, Tatsuo Mano, Yoshiki Niimi, Atsushi Iwata, Tatsushi Toda, Takeshi Iwatsubo

## Abstract

**Background:** The COVID-19 pandemic has affected not only the emergency medical system, but also patients’ regular ambulatory care. The number of patients visiting outpatient internal medicine clinics decreased during March–April 2020 compared to 2019. Moreover, the ban on telephone re-examination for outpatient clinics in lieu of ambulatory care for chronic diseases has been lifted since March 2020. In this context, we investigate the impact of the COVID-19 pandemic on ambulatory care at Japanese outpatient clinics for patients with chronic neurological diseases during the first half of 2020.

**Methods:** We collected data from the administrative claims database by DeSC Healthcare. Serial changes in the frequency of subsequent outpatient visits to clinics or hospitals (excluding large hospitals with beds >200) for chronic ambulatory care of epilepsy, migraine, Parkinson’s disease (PD), and Alzheimer’s disease were measured. We also evaluated the utilization rate of telephone re-examination at outpatient clinics.

**Results:** Since April 2020, the monthly count of outpatient clinic visits for epilepsy or PD decreased slightly but significantly. The use of telephone re-examination was facility-dependent, and it was used in less than 5% of all outpatient clinic visits for the examined neurological diseases in May 2020. The utilization rate of telephone re-examination was not associated with age or the neurological diseases of interest.

**Conclusion:** The impact of the COVID-19 pandemic on ambulatory care for several chronic neurological diseases may have been relatively limited, in terms of the frequency or type of outpatient visit, during the first half of 2020 in Japan.

## 1. Introduction

The global coronavirus disease (COVID-19) pandemic, since early 2020, has severely affected not only the emergency medical system in Japan [1], but also patient care at the outpatient clinics. To reduce the risk of COVID-19 infection, people were requested to refrain from unnecessary and nonurgent outings or from visiting crowded places, with a call to “avoid the three Cs” (closed spaces, crowded places, and close-contact settings) [2] based on the Act on Special Measures for Pandemic Influenza and New Infectious Diseases Preparedness and Response (https://elaws.e-gov.go.jp/document?lawid=424AC0000000031). People also refrained from visiting outpatient clinics [3], leading to a significant decrease in the number of ambulatory visits to internal medicine outpatient clinics in Japan by more than 10% in April–May 2020 compared to April–May 2019 [4]. In addition, the ban on telephone re-examination at outpatient clinics as an alternative to ambulatory care for chronic diseases was removed by the Ministry of Health, Labor and Welfare (MHLW) since March 2020 as an exceptional measure against the COVID-19 pandemic [5]. Specifically, in terms of reimbursement, it became newly available to claim a “subsequent visit fee” along with the “prescription fee,” even in the case of telephone re-examination.

As patients with chronic neurological diseases need continuous medication and regular re-examination at the outpatient clinic, it is unfavorable to interrupt ambulatory care completely, even during a state of emergency [6]. In this context, we infer that some patients with neurological disease may have adapted by increasing the number of prescription days or by using telephone re-examination [7, 8], thereby attempting to decrease the frequency of direct visits to the outpatient clinic [9]. However, despite such modest responses on the part of the patient, there may remain some medium- or long-term safety concerns about their disease control, because in-person re-examination is deemed essential, especially for the ambulatory care of patients with neurological diseases [10].

Although an earlier questionnaire survey administered across hundreds of clinics and hospitals in Japan [4] reported that telephone re-examination was used only in a small proportion of outpatient visits (i.e., in approximately 2% of all subsequent visits), safety and efficacy of implementing telephone re-examination as one of the temporary measures under the COVID-19 pandemic has hardly been evaluated. Much less, in the first place, evidence is generally limited on the impact of the COVID-19 pandemic on the care for patients with chronic neurological diseases in Japan [11]. Therefore, we aimed to assess the basic features of the change in care for patients with chronic neurological disease at the outpatient clinic during the first half of 2020. We used the DeSC claims database that is based on the Japanese public health insurance and comprises data on more than one million individuals in Japan. The database has a great advantage in terms of its high accessibility and analyzability compared to much larger Japanese claims databases (e.g., NDB) [12], and thus, the use of the DeSC database in the current study would provide a starting point and an important foundation for further research.

## 2. Materials and Methods

### 2.1. Study design

This was a retrospective observational study using administrative claims data and was approved by the University of Tokyo Graduate School of Medicine Institutional Ethics Committee (ID: 11628-(3)). Informed consent is not required because this study only uses already-prepared anonymized information as required by the Act on the Protection of Personal Information in Japan (https://elaws.e-gov.go.jp/document?lawid=415AC0000000057). We applied for access to the DeSC database (https://desc-hc.co.jp/archives/2188) in March 2021, which was approved by DeSC Healthcare, Inc. (https://desc-hc.co.jp/en), permitting us to use the data in April 2021.

### 2.2. About the DeSC database

The DeSC database was built by anonymizing and processing data from the health insurance claims database provided by several Japanese public health insurers covering more than one million individuals. Two types of insurers are included: National Health Insurance (NHI) and society-managed, employment-based health insurance association (SHI) [13]. The details of these health insurers, including their names or addresses, are completely confidential. The degree of overlap in the geographical medical areas of NHI and SHI insurers is also undisclosed. The eligibility criteria for the DeSC database is as follows: insured individuals and their dependents who are in the age group of 19–74 years as on June 30, 2021, in SHI, or those who are in the age group of 0–74 years on July 31, 2021, in NHI. The database includes all eligible patients’ receipts claimed during the period between April 2015 and June 2021 (in SHI) or the period from April 2015 to July 2021 (in NHI). Other types of Japanese health insurers covering those younger than 75 years old, such as public-corporation-run health insurance or mutual aid association, are not included in this database. In addition, in Japan, people who are 75 years or older are covered by other specific health insurance schemes such as “Late-stage medical care system for the elderly aged 75 and over” [13], and thus, older adults (75 years and above) are not included in this database.

As for database specifications, disease name, drugs, or medical procedures related to the following specific diseases are masked for anonymity: “designated infectious disease” such as COVID-19 or “type 1-3 infectious diseases” (e.g., Ebola hemorrhagic fever, tuberculosis, SARS, or cholera) as specified in the Act on the Prevention of Infectious Diseases and Medical Care for Patients with Infectious Diseases in Japan (https://elaws.e-gov.go.jp/document?lawid=410AC0000000114), or “designated intractable diseases” [14] excluding Parkinson’s disease or ulcerative colitis.

### 2.3. Data preprocessing

The process of data handling and analyses were conducted using R software (version 3.5.1) in MacOS Catalina. The preprocessing workflow is shown in Fig. 1. First, the total number of insured individuals and their dependents listed by the insurers, during the study period from April 2015 to June 2020 in SHI (or July 2020 in NHI) were 682,186 in SHI and 603,798 in NHI (excluding those who had withdrawn from the insurers’ list during the study period, or those data which encountered some data processing failure during the database formation).

**Figure 1.**
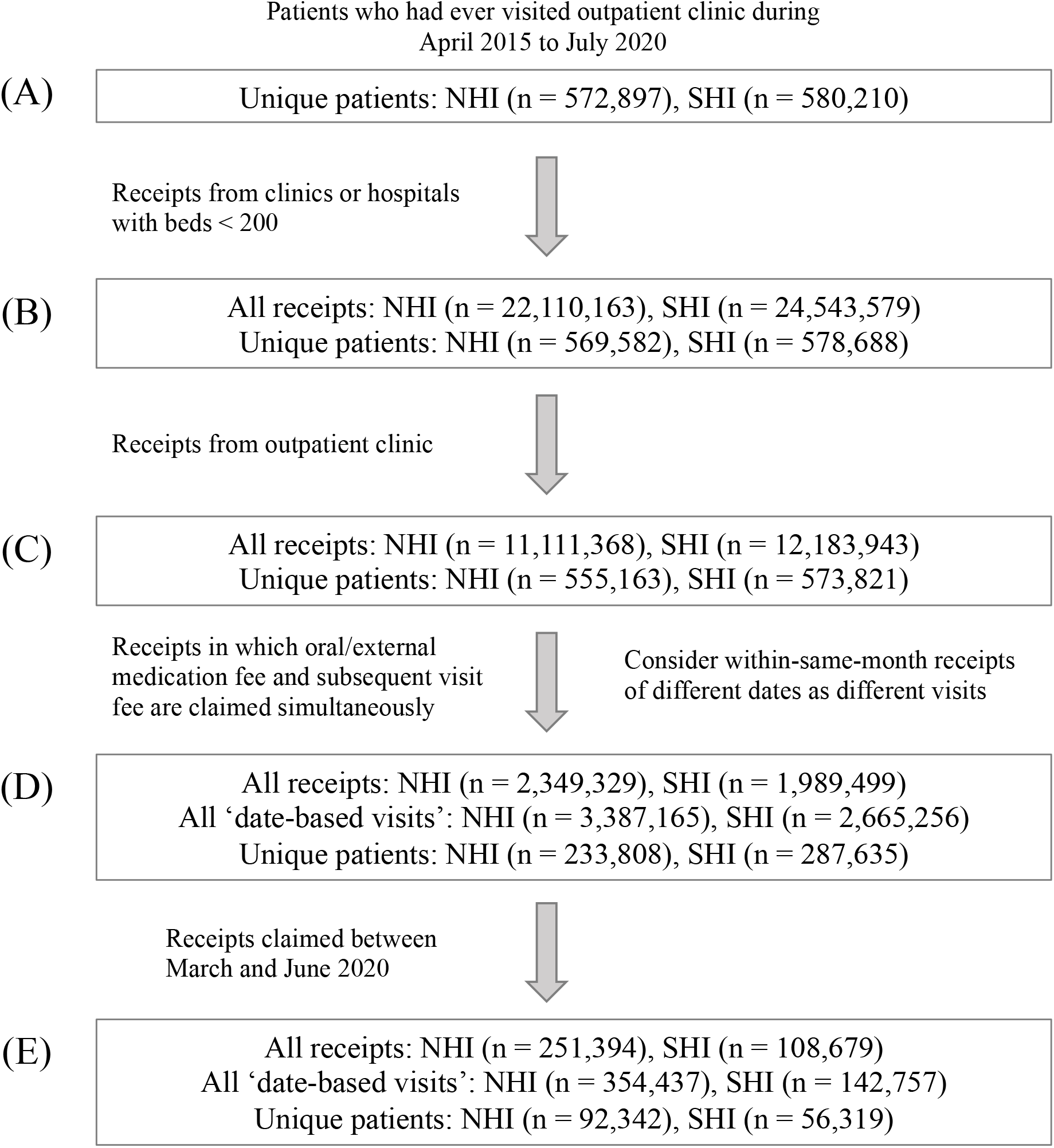
Data preprocessing workflow Abbreviations: SHI, society-managed, employment-based health insurance association; NHI, National Health Insurance

Among them, there were 580,210 unique patients in SHI and 572,897 unique patients in NHI who had ever visited outpatient clinic during the study period of April 2015 to June 2020 in SHI (or July 2020 in NHI) (Figure 1A). Receipts were claimed for each patient in each month of visit by each clinic or hospital, and for each type of medical setting (e.g., ambulatory care, in-hospital care, or pharmacy); for our analysis, we only included the receipts claimed for ambulatory care at the facilities with less than 200 beds (Figure 1B). We limited the size of clinics or hospitals of each receipt, because re-examination at facilities with 200 or more beds are claimed under “outpatient clinic fee” (medical practice code: 112011310) regardless of the use of telephone or any online devices, and consequently, the use of telephones for re-examination cannot be identified from the data. In the case of facilities with less than 200 beds, ambulatory care via the telephone or any electronic device can be identified, since it would be claimed as a “subsequent visit fee by telephone” (medical practice code (version in April 2020): 112007950, 112008850, and 112023350) instead of simply “subsequent visit fee” (medical practice code: 112007410, 112015810, and 112008350). We have not considered the claims under “first visit fee” (medical practice code: 111000110) or “online medicine fee” (medical practice code: 112023210), because in case of the former, one cannot distinguish whether the examination was done by in person or via telephonic examination, and in the case of the latter claims, facilities are required to notify authorities in advance to start “online medicine,” which have actually been hardly used in this database even during the COVID-19 pandemic.

Subsequently, among the receipts claimed for ambulatory care at the outpatient clinic (Figure 1C), we considered only those that included the prescription of oral or external medications as well as the claims of “subsequent visit fee” or “subsequent visit fee by telephone” for the analysis (Figure 1D). Next, since different outpatient visits within the same month cannot be distinguished solely by each receipt, we additionally referred to the date of visits to differentiate between each visit. We use the term “date-based visit” to refer to the distinguished minimum unit of visits and conducted a visit-based analysis.

### 2.4. Outcome definition

For the above-mentioned date-based visits, we determined the diseases for which each patient visited the outpatient clinic. Here, we focus on several common neurological diseases, including Alzheimer’s disease (AD) dementia, epilepsy, Parkinson’s disease (PD), and migraine. Rarer neurological diseases such as spinocerebellar degeneration, myasthenia gravis, or multiple sclerosis cannot be identified because they have been anonymized during the database creation process. We did not include chronic ischemic stroke or chronic hemorrhagic stroke because they cannot always be specifically identified by the name of the medication. In addition, we also focused on the popular risk factors for vascular diseases, namely hypertension (HTN), diabetes (DM), and dyslipidemia (DL).

We determined whether each date-based outpatient visit was related to the care of each disease, defined by fulfilling both of the following criteria: (A) medications specific to the disease of interest are prescribed (Table 1A), and (B) receipt claim (monthly) includes disease names related to the disease of interest (Table 1B). This is in reference to earlier studies (e.g., in PD [15, 16] or epilepsy [17, 18]). We referred to the Anatomical Therapeutic Chemical classification system (ATC code) for identifying medications (A) and the 10th version of the International Statistical Classification of Diseases and Related Health Problems (ICD-10) for identifying diseases (B).

**Table 1.** Disease definition for the date-based outpatient clinic visits

| Disease | (A) by diagnosis (ICD-10 subcode) | (B) by medication (ATC subcode) |
| --- | --- | --- |
| Alzheimer's disease | G30, G31, F00, F01, F02, F03 | N07D0 |
| Epilepsy | G40 | N03A0 |
| Parkinson's disease | G20 | N04A0 |
| Migraine | G43 | N02C0 |
| Hypertension | I10, I11, I12, I13 | C08, C09 |
| Diabetes | E10, E11, E12, E13, E14, R73 | A10 |
| Dyslipidemia | E78 | B01 |

Abbreviations: ICD-10, 10th version of the International Statistical Classification of Diseases and Related Health
Problem; ATC code, Anatomical Therapeutic Chemical Classification System.

### 2.5. Statistical analyses

The DeSC database comprises two sub-datasets (i.e., the NHI and SHI datasets), corresponding to data obtained from different types of insurers. The population covered by these insurers have different disease frequencies partly due to the differences in age distribution (as mentioned in the Results section) and social background (e.g., SHI is for those working at certain companies, but NHI is not limited to active workers). Therefore, we conducted the same analysis on each of the two datasets independently, and then comparatively described the results. In this study, we performed the following analyses.

1. Serial monthly changes in the date-based visit of neurological diseases of interest, in terms of before and after the nationwide declaration of a state of emergency [6] in early April 2020.
2. Facility-dependent variance in the utilization (frequency or rate) of telephone re-examination.
3. Utilization rate of telephone re-examination among date-based outpatient visits since March 2020.
4. Features which promoted or discouraged the use of telephone re-examination.

First, we evaluated the overlap in insurers (i.e., NHI or SHI here) among all visits in each facility, because the degree of overlap in the geographical medical areas of NHI and SHI insurers is uncertain. In Japan there is no restriction on patients regarding which hospitals to visit (“free access”) regardless of the type of health insurance; in this case, if many facilities had received only the patients covered by one type of insurer, the locations of the NHI and SHI insurers should be interpreted as geographically located away from each other. Such insurer-deviation in each facility *F* is evaluated by the asymmetry index (AI), calculated by the following formula:

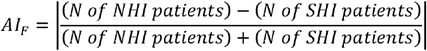

where AI is in the range of [0-1]. If AI is not 1 in many of the facilities, the residing locations of patients insured by NHI and SHI can then be interpreted as geographically overlapping with each other.

For Analysis (1), serial change at each month since the nationwide declaration of a state of emergency (in early April 2020) in comparison with the previous year was measured by relative risk (RR), calculated by the following formula:

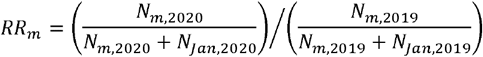

Where *N*_*m*,2020_ denotes the total number of date-based visits of a certain disease of interest in month *m* of 2020. As a reference, we used the total number of date-based visits in January 2019 and January 2020. The minimum value among *RR*_*Mar*_ through *RR*_*Jun*_ was chosen to measure the largest degree of decrease in monthly outpatient visits. When the upper 95% of the selected RR was less than 1, or when the lower 95% of the selected RR was larger than 1, the number of date-based visits was considered to have significantly changed compared to the previous year. To calculate RR and its 95% CI, we used the R package {*epitools*} [19].

In addition, we also applied interrupted time-series analysis (ITSA) [20] for Analysis (1), based on the impact model where a decline in the level occurs along with the declaration of a state of emergency (in April 2020), but the long-term serial trend was maintained. The Poisson regression formula for the ITSA is as follows:

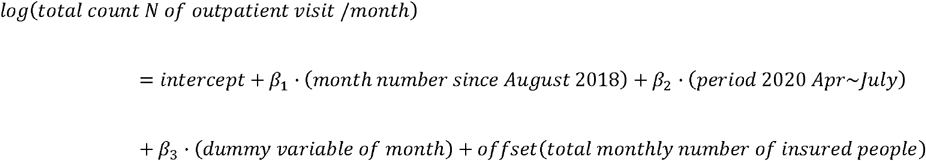

where the term “*month number since August* 2018” denotes long-term trend (slope) of the month since August 2018, and the term “*Period* 2020 *Apr ∼ July*” means that it is after the declaration of state of emergency. Seasonality in the serial change of monthly outpatient clinic visits is taken into account by the term “*dummy variable of month*”. The visit count is also influenced by the total number of insured individuals and is thus considered as the offset term. As in RR, when the upper 95% confidence interval (CI) of the adjusted odds ratio (OR) was less than 1, or when the lower 95% CI of the adjusted OR was larger than 1, the factor was considered to be significantly associated with the utilization of telephone re-examination in the regular outpatient visit.

In Analysis (2), we calculated the coefficient of variation (*CV*_*D*_) of each disease *D* to measure the facility-dependent variance in the utilization rate of telephone re-examination between March and June 2020, using the following formula:

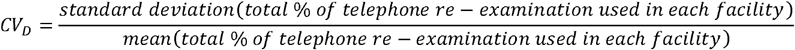

We regarded the use of telephone re-examination for disease *D* to vary based on the facility if CV_D_ > 1. We excluded facilities that had never claimed telephone re-examination between March and June 2020 in this calculation.

In Analysis (4), we merged NHI and SHI datasets together, and applied a generalized linear mixed model to examine which features are associated with the use of telephone re-examination among all the date-based outpatient clinic visits. Since there is high variance in the number of telephone re-examination depending on the clinic or hospital (as observed in Analysis (2)), as well as differences between the two datasets in terms of age distribution and disease frequency, we incorporated the factor of patient care facility and the insurer of each patient into the mixed effects model as random effect terms. The model is described by the following formula [21]:

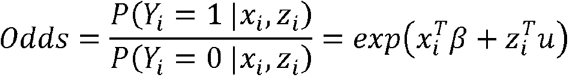

where *Y*_*i*_ represents the binomial status of the use or non-use of telephone re-examination in the date-based outpatient clinic visits *i* for any diseases, *β* is the vector of fixed effect parameters, *x*_*i*_ is the covariate matrix for fixed effects, *u* is the vector of random effect parameters, and *z*_*i*_ is the covariate matrix of random effects. Covariates to measure fixed effects include patient’s age at the time of outpatient clinic visit, sex of the patient, and binomial status whether the visit is for seeking care for a certain disease of interest. Covariates used to measure random effects (as the random intercept) denote the factor of the clinic or hospital where the receipt was claimed, and the type of insurance (i.e., NHI or SHI). For the multilevel analysis, we used the R package {*lme4*} [22].

## 3. Results

### 3.1. Basic demographic characteristics

We included 2,665,256 date-based outpatient clinic visits claimed during the study period from 287,635 unique patients in SHI and 3,387,165 date-based visits claimed during the study period from 233,808 unique patients in NHI (Figure 1D). As expected, AI was mostly 1 in many facilities (mean = 0.977, 95%-tile = 1.000, and 99.7%-tile = 1.000).

The summary demographic characteristics of eligible date-based visits and unique patients is described in Table 2. When categorized by visits (Table 2A), the median age of patients covered by SHI was 51.7 years (IQR: 42.3-58.7) and 69.0 years (IQR: 61.5-72.1) for those covered by NHI, revealing a clear difference between the two insurers. The distribution of patients in terms of sex was mostly equivalent; 49.2% in SHI and 53.7% in NHI were female. The female patients were slightly younger than the male patients in both SHI and NHI (by approximately 1–2 years). Among all eligible date-based visits, telephone re-examination was used in 0.09% (2,279/2,665,256) in SHI and 0.1% (3,259/3,387,165) in NHI. In addition, among the eligible visits from March to June 2020, telephone re-examination was used in 0.34% (479/142,757) in SHI and 0.57% (2,555/446,727) in NHI.

**Table 2.** Basic demographic characteristics of all eligible outpatient visits / patients from NHI and SHI

| (A) Summary, by visit |  | SHI |  | NHI |  |
| --- | --- | --- | --- | --- | --- |
|  |  | All visits | Visits during Mar–Jun 2020 | All visits | Visits during Mar–Jul 2020 |
| N |  | 2665256 | 142757 | 3387165<br>( 100 % ) | 354437 ( 100 % ) |
| Age (in Aug 2020) |  | 51.7 ( 42.3 ~ 58.7 ) | 51.6 ( 42.4 ~ 58.1 ) | 69 ( 61.5 ~ 72.1 ) | 69 ( 62.6 ~ 72.1 ) |
| Sex (female) |  | 1309885<br>( 49.15 % ) | 67598 ( 47.35 % ) | 1820169<br>( 53.74 % ) | 191812 ( 54.12 % ) |
| Telephone re-examination used |  | 2279 ( 0.09 % ) | 479 ( 0.34 % ) | 3259 ( 0.1 % ) | 2555 ( 0.57 % ) |
| Neurological diseases | Alzheimer's disease | 291 ( 0.01 % ) | 10 ( 0.01 % ) | 4275 ( 0.13 % ) | 624 ( 0.18 % ) |
|  | Epilepsy | 8640 ( 0.32 % ) | 490 ( 0.34 % ) | 39956 ( 1.18 % ) | 3667 ( 1.03 % ) |
|  | Parkinson's disease | 1879 ( 0.07 % ) | 103 ( 0.07 % ) | 30307 ( 0.89 % ) | 2347 ( 0.66 % ) |
|  | Migraine | 7567 ( 0.28 % ) | 410 ( 0.29 % ) | 5649 ( 0.17 % ) | 538 ( 0.15 % ) |
| Other diseases for reference | Hypertension | 336165<br>( 12.61 % ) | 21586 ( 15.12 % ) | 860634<br>( 25.41 % ) | 98302 ( 27.73 % ) |
|  | Diabetes | 89635 ( 3.36 % ) | 5717 ( 4 % ) | 213799<br>( 6.31 % ) | 24757 ( 6.98 % ) |
|  | Dyslipidemia | 254915<br>( 9.56 % ) | 16896 ( 11.84 % ) | 559083<br>( 16.51 % ) | 67105 ( 18.93 % ) |

| (B) Summary, by unique patients |  | SHI |  | NHI |  |
| --- | --- | --- | --- | --- | --- |
|  |  | All patients | Patients who visited during Mar–Jun 2020 | All patients | Patients who visited during Mar–Jun 2020 |
| N |  | 287635 | 56319 | 233808 | 92342 |
| Age (in Aug 2020) |  | 46.6 ( 36.1 ~ 55.1 ) | 49.6 ( 39.9 ~ 56.9 ) | 66.6 ( 49.8 ~ 71.4 ) | 68.8 ( 61.5 ~ 72 ) |
| Sex (female) |  | 141495<br>( 49.19 % ) | 27401 ( 48.65 % ) | 131095<br>( 56.07 % ) | 52219 ( 56.55 % ) |
| Telephone re-examined experienced |  | 1938 ( 0.67 % ) | 441 ( 0.78 % ) | 2089 ( 0.89 % ) | 1423 ( 1.54 % ) |
| Neurological diseases | Alzheimer's disease | 23 ( 0.01 % ) | 6 ( 0.01 % ) | 334 ( 0.14 % ) | 188 ( 0.2 % ) |
|  | Epilepsy | 493 ( 0.17 % ) | 169 ( 0.3 % ) | 1687 ( 0.72 % ) | 1009 ( 1.09 % ) |
|  | Parkinson's disease | 123 ( 0.04 % ) | 28 ( 0.05 % ) | 909 ( 0.39 % ) | 531 ( 0.58 % ) |
|  | Migraine | 1164 ( 0.4 % ) | 209 ( 0.37 % ) | 643 ( 0.28 % ) | 187 ( 0.2 % ) |
| Other diseases for reference | Hypertension | 14257 ( 4.96 % ) | 7714 ( 13.7 % ) | 39570<br>( 16.92 % ) | 28651 ( 31.03 % ) |
|  | Diabetes | 3544 ( 1.23 % ) | 1898 ( 3.37 % ) | 10152 ( 4.34 % ) | 7039 ( 7.62 % ) |
|  | Dyslipidemia | 12541 ( 4.36 % ) | 6344 ( 11.26 % ) | 27850<br>( 11.91 % ) | 19923 ( 21.58 % ) |

Abbreviations: SHI, society-managed, employment-based health insurance association; NHI, National Health
Insurance

### 3.2. Serial changes in outpatient clinic visits

First, we reviewed serial changes in the date-based outpatient clinic visits within the last two years, as shown in Figure 2. Visual inspection revealed that the number of monthly visits for any disease (Figure 2A), for hypertension (HTN) or dyslipidemia (DL) (Figure 2B), or for epilepsy or PD (Figure 2C) appeared to be decreasing slightly since April 2020.

**Figure 2.**
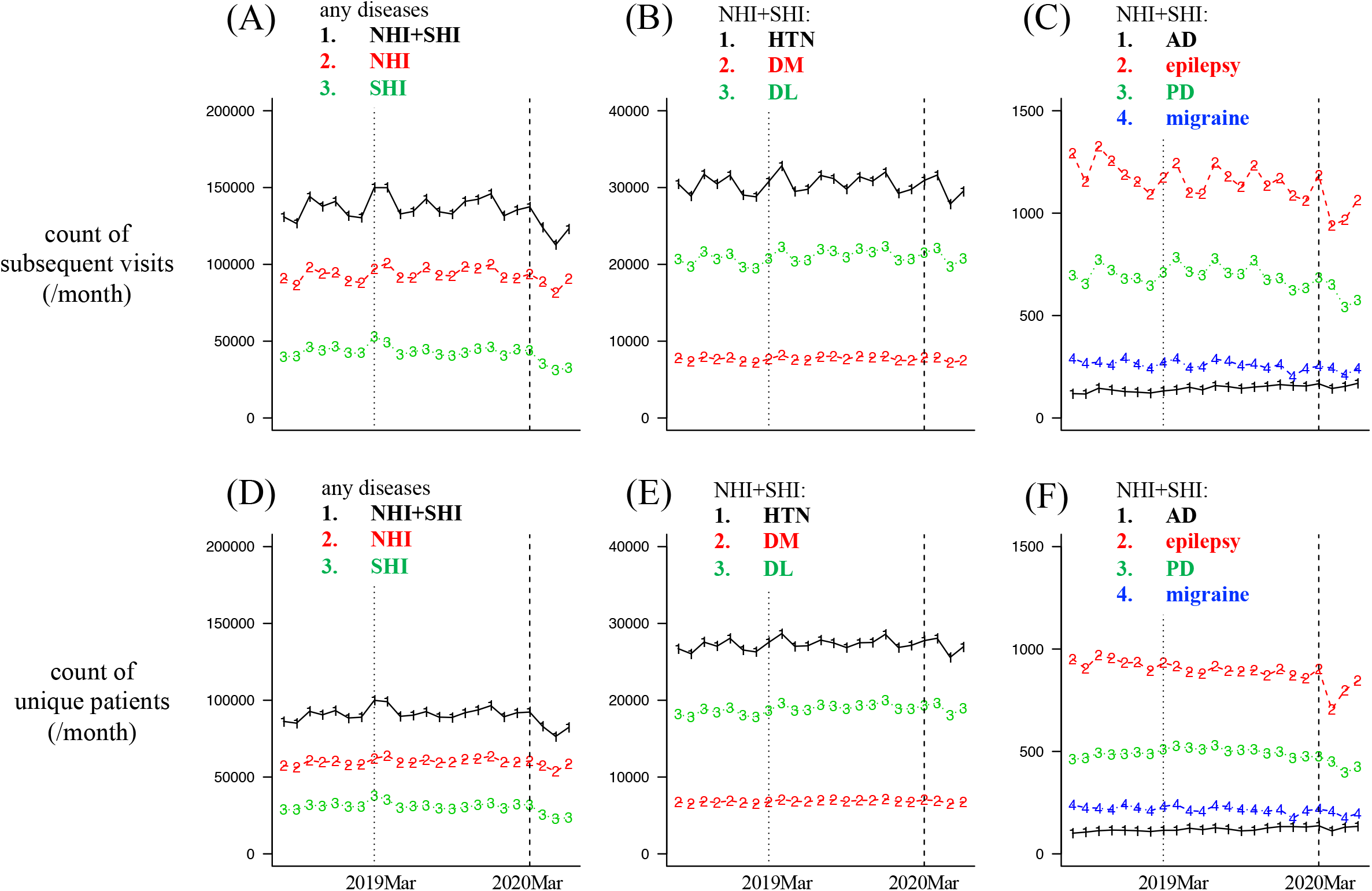
Serial change in the monthly count of visits / patients where subsequent visit fee were claimed, since August 2018 to June 2020 The vertical dashed line shows March 2020, and the vertical dotted line indicates March 2019. Abbreviations: SHI, society-managed, employment-based health insurance association; NHI, National Health Insurance; AD, Alzheimer’s disease; PD, Parkinson’s disease; HTN, hypertension; DM, diabetes; DL, dyslipidemia

This was statistically validated by RR and ITSA, showing similar results (Table 3): in NHI, the number of monthly visits for epilepsy, PD, HTN, DM, and DL significantly decreased in both RR (Table 3A) and ITSA (Table 3B). Meanwhile, in SHI, the number of monthly visits for HTN, DM, and DL, but not for neurological diseases, significantly decreased in both RR (Table 3A) and ITSA (Table 3B). Regular outpatient visits (with “subsequent visit fee” claimed) for any diseases decreased with an RR of 0.87 (95% CI: 0.86 ∼ 0.88) in SHI and RR = 0.93 (95% CI: 0.93 ∼ 0.94) in NHI (Table 3A), which is consistent with the earlier reports by Japanese Medical Associations published in June 2020 [4]. Moreover, we observed similar results when conducting the same analysis based on the monthly count of unique patients for each disease (Supplemental Table 1).

**Table 3.** Decrease in the number of outpatient visits since April 2020: RR and ITSA

( A ). RR results and 95% CI
| (by visits)<br>Disease | SHI<br>minimum RR | NHI<br>minimum RR |
| --- | --- | --- |
| Alzheimer's disease | 0.476 ( 0.133 ~ 1.706 ) , May | 0.91 ( 0.772 ~ 1.073 ) , April |
| Epilepsy | 1.021 ( 0.85 ~ 1.228 ) , June | 0.872 ( 0.817 ~ 0.93 ) * , April |
| Parkinson's disease | 0.776 ( 0.514 ~ 1.173 ) , June | 0.905 ( 0.833 ~ 0.982 ) * , May |
| Migraine | 1.038 ( 0.875 ~ 1.231 ) , March | 0.99 ( 0.846 ~ 1.16 ) , April |
| Hypertension | 0.925 ( 0.901 ~ 0.951 ) * , June | 0.967 ( 0.955 ~ 0.98 ) * , May |
| Diabetes | 0.897 ( 0.851 ~ 0.946 ) * , June | 0.964 ( 0.94 ~ 0.989 ) * , May |
| Dyslipidemia | 0.924 ( 0.896 ~ 0.953 ) * , June | 0.962 ( 0.948 ~ 0.977 ) * , May |
| any diseases | 0.868 ( 0.859 ~ 0.876 ) * , April | 0.93 ( 0.924 ~ 0.936 ) * , April |

( B ). Results of ITSA as an event intercept and its 95% CI
| (by visits)<br>Disease | SHI<br>Impact in April 2020 | NHI<br>Impact in April 2020 |
| --- | --- | --- |
| Alzheimer's disease | 0.474 ( 0.13 ~ 1.623 ) | 0.893 ( 0.775 ~ 1.029 ) |
| Epilepsy | 1.091 ( 0.922 ~ 1.291 ) | 0.909 ( 0.862 ~ 0.96 ) * |
| Parkinson's Disease | 0.703 ( 0.492 ~ 1.001 ) | 0.816 ( 0.763 ~ 0.873 ) * |
| Migraine | 1.057 ( 0.883 ~ 1.266 ) | 0.895 ( 0.776 ~ 1.032 ) |
| Hypertension | 0.896 ( 0.874 ~ 0.919 ) * | 0.965 ( 0.955 ~ 0.976 ) * |
| Diabetes | 0.882 ( 0.84 ~ 0.926 ) * | 0.961 ( 0.941 ~ 0.982 ) * |
| Dyslipidemia | 0.891 ( 0.866 ~ 0.916 ) * | 0.96 ( 0.947 ~ 0.972 ) * |
| Any diseases | 0.762 ( 0.755 ~ 0.769 ) * | 0.903 ( 0.898 ~ 0.908 ) * |

Abbreviations: RR, relative risk; ITSA, interrupted time-series analysis; CI, confidence interval; SHI,
society-managed, employment-based health insurance association; NHI, National Health Insurance.

### 3.3. Facility-dependent variability in telephone re-examination use

Whether telephone re-examination is used during regular visits depends not only on the patients’ intention but also on the status of each facility (clinic/hospital) to enable the use of telephone re-examination, as well as any compelling need to use telephone re-examination, such as forced temporary closure of outpatient clinics due to the nosocomial outbreak of COVID-19 [23]. This means that the utilization rate varies depending on each facility.

To measure the degree of such facility-dependent variability in the use of telephone re-examination, we first calculated the CV of the telephone re-examination rate. The results revealed an especially high variance for epilepsy, although the facility-dependent variance was generally mildly high (CV > 1): CV_AD_ = 1.60, CV_epilepsy_ = 2.73, CV_PD_ = 2.29, CV_migraine_ = 2.22, CV_HTN_ = 1.29, CV_DM_ = 1.35, CV_DL_ = 1.27, and CV_any_ = 1.36 (Figure 3A).

**Figure 3.**
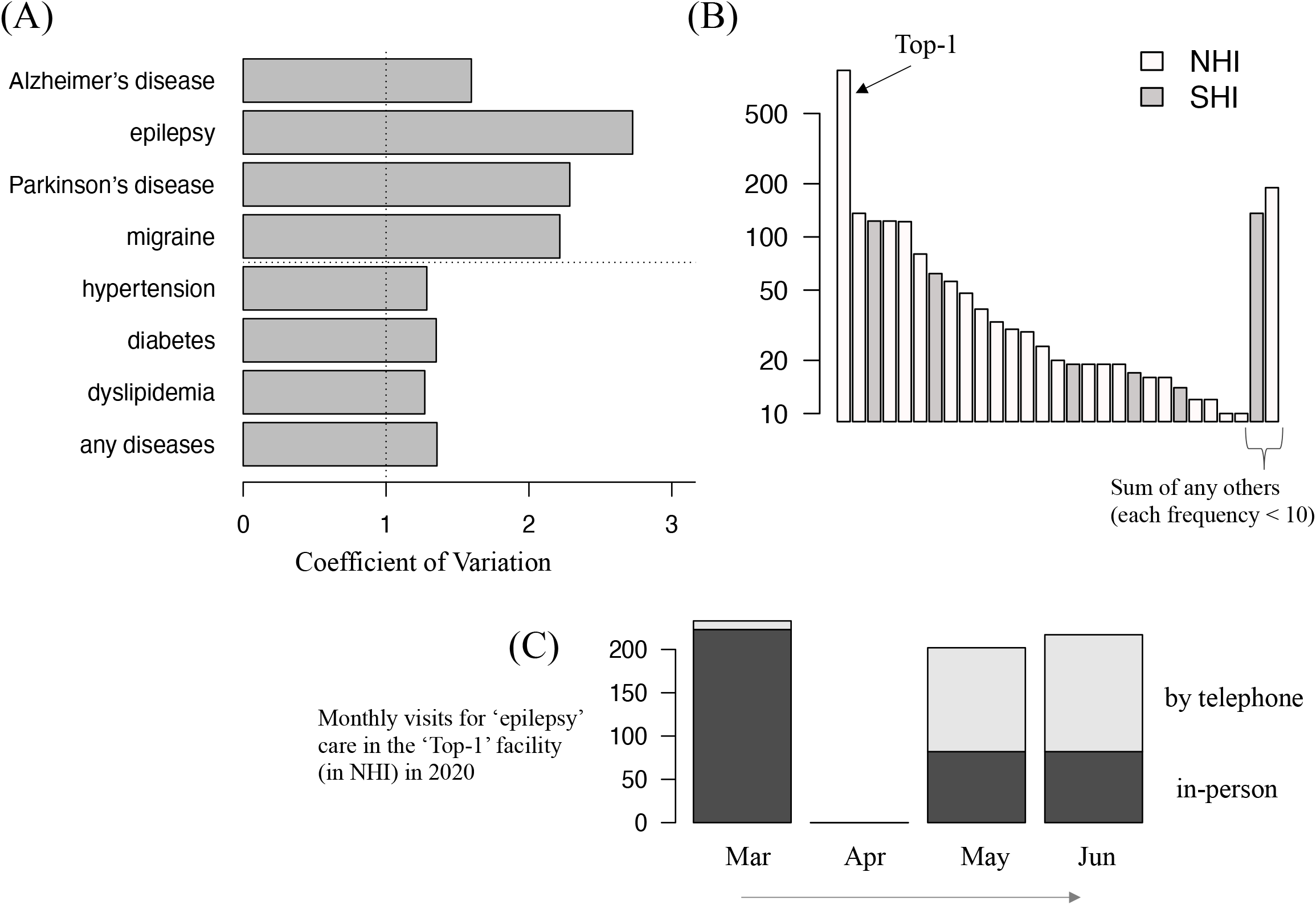
Facility-dependent variance in the number/rate of telephone re-examination used in subsequent outpatient visits between March and June 2020 (A) Visits for epilepsy had the highest CV (= 2.73), while the utilization of telephone re-examination is generally facility-dependent (CV for any diseases = 1.36). (B) Bar plot of ambulatory care visits by telephone re-examination at all facilities between March and June 2020, regardless of the type of insurance. (C) Serial changes in the number of visits for ambulatory care for epilepsy at the ‘Top-1’ outpatient clinic. Abbreviations: SHI, society-managed, employment-based health insurance association; NHI, National Health Insurance

Figure 3B is a bar plot showing the number of outpatient clinic visits by facility (clinics or hospitals) where telephone re-examination “subsequent visit fee” had been claimed from March 2020 to June 2020 for 10 or more visits (NHI and SHI visits were separately counted in each facility). A large variability was observed, and the “Top-1” facility accounted for 35.9% of all telephone re-examination visits since March 2020.

Notably, when we focused on the “Top-1” facility in NHI in terms of its care for epilepsy (Figure 3C), there were no epilepsy patients’ ambulatory care visits in April 2020, and subsequently, the proportion of telephone re-examination use clearly increased in May and June 2020.

### 3.4. Telephone re-examination utilization

Serial changes in the utilization rate of telephone re-examination at outpatient clinics are plotted in Figure 4. In SHI (Figure 4A-C), among the neurological diseases of interest, telephone re-examination was most frequently used in visits for migraine (4.44%) in May 2020 (Figure 4A), followed by visits for epilepsy (1.72%). Further, visits for HTN, DM, or DL had an approximately 2% utilization rate of telephone re-examination in May 2020 (Figure 4C). On the other hand, in NHI, telephone re-examination was most frequently used in visits for epilepsy among the neurological diseases of interest (Figure 4D): 14.4% in May 2020. Meanwhile, HTN, DM, and DL in NHI had a utilization rate of approximately 1% or less in May 2020 (Figure 4F).

**Figure 4.**
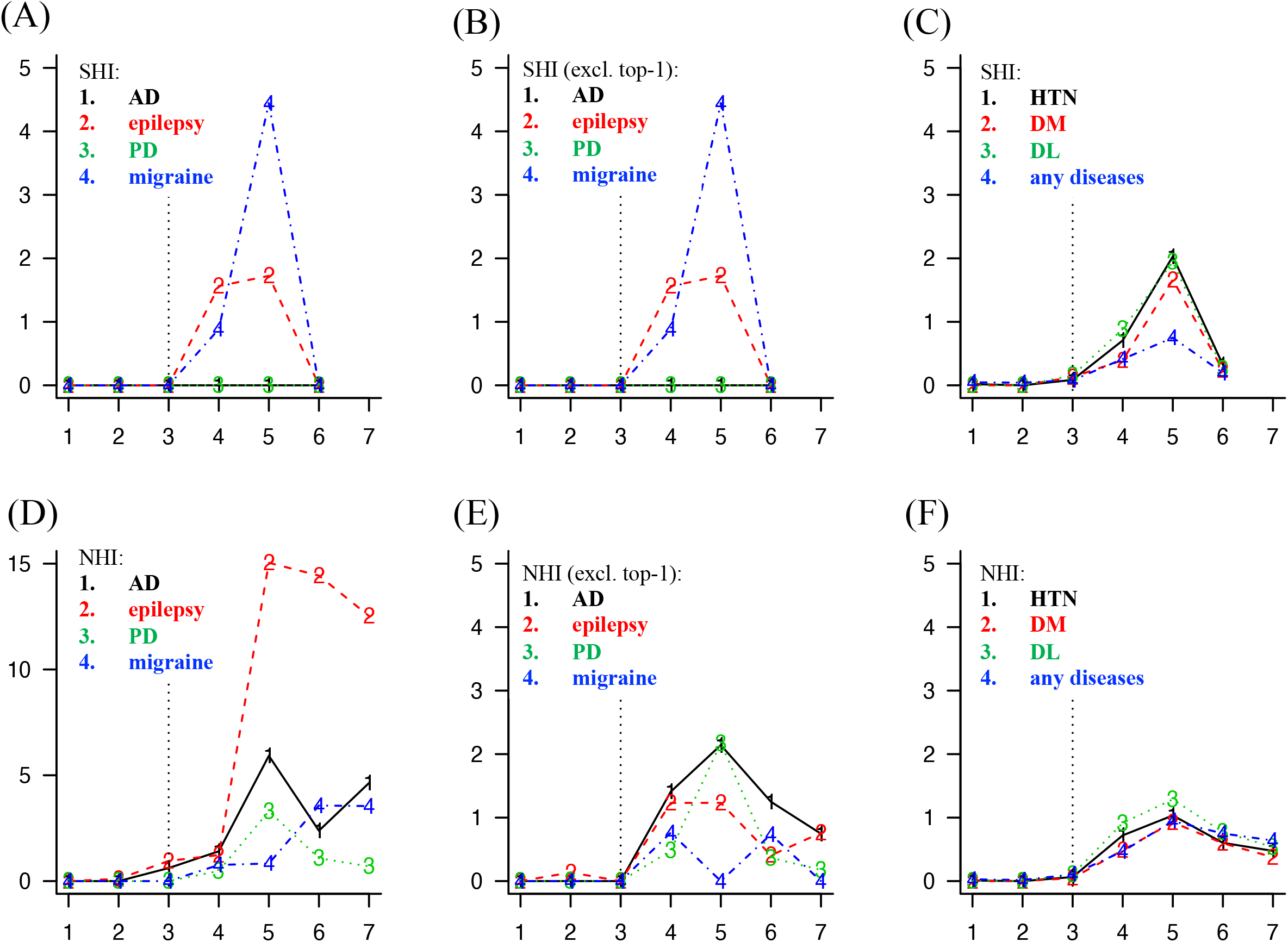
Serial increase in the monthly utilization rate (%) of telephone re-examination among all types of re-examination visits at outpatient clinic Serial changes between January and July 2020. The vertical dotted line represents March 2020. Please note that the eligible period is until June 2020 in SHI (A-C) and until July 2020 in NHI (D-F). In addition, the y-axis scale is 0-15 only in (D), whereas it is 0-5 in others. Since there were no visits to the ‘Top-1’ facility by patients insured with the SHI, (A) and (B) are the same plots. Abbreviations: SHI, society-managed, employment-based health insurance association; NHI, National Health Insurance; AD, Alzheimer’s disease; PD, Parkinson’s disease; HTN, hypertension; DM, diabetes; DL, dyslipidemia

Since there was variability in the use of telephone re-examination depending on each facility (e.g., “Top-1” facility in Figure 3B), we excluded visits from the “Top-1” facility (in NHI) and plotted the same serial changes in Figures 4B and 4E. In NHI excluding its “Top-1” facility (Figure 4E), telephone re-examination was used in visits for epilepsy (constituting less than 2%) in May 2020, and in visits for AD or PD (approximately 2%) in May 2020.

### 3.5. Factors associated with the use of telephone re-examination

We shall now examine which features of each visit may have contributed (or not contributed) to the use of telephone re-examination in regular visits. The results are shown in Table 4, where the values in each cell denote the fixed effect coefficient and its 95% CI of the covariate in the same row for the target variable in the same column. After adjusting for the type of insurers and the facility-specific factor, female patients were found to be slightly more likely to use telephone re-examination than men (adjusted OR: approximately 1.3) consistently, while patient age was not associated with the use of telephone re-examination. Regular visits for neurological diseases of interest were not associated with the use of telephone re-examination. Interestingly, regular visits for HTN or DL were inclined to be conducted via telephone (adjusted OR: approximately 1.3), while the visits for DM were less likely to be telephonic (adjusted OR: 0.64). The same analysis was not conducted for the patient-based count table (Table 2B) because of the difficulty in adjusting the factors influencing facility visits (e.g., some patients regularly visit several different facilities).

**Table 4.**
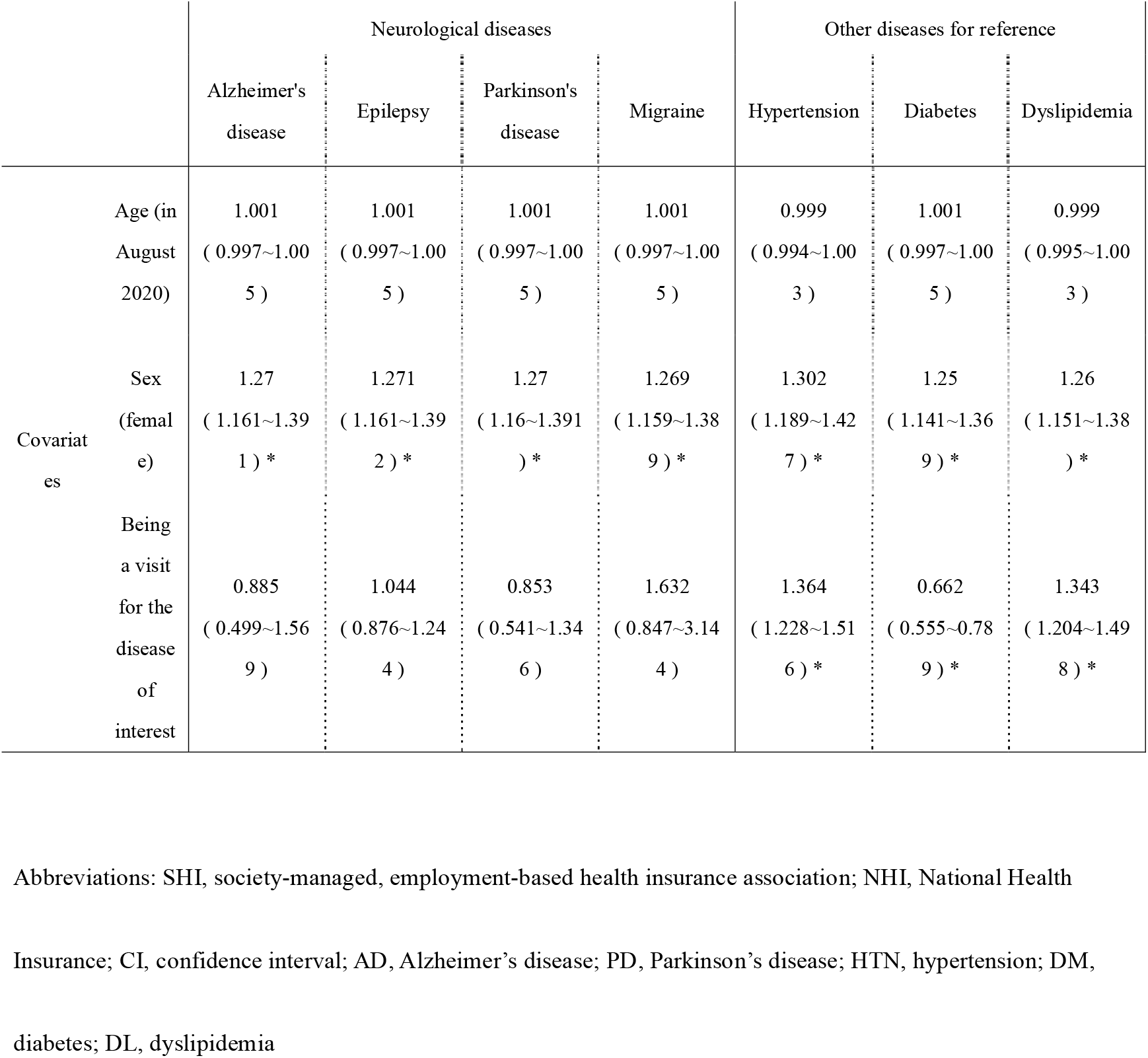
Results of the generalized linear mixed model for all (SHI + NHI) receipts between March and June 2020: Fixed-effects coefficients and their 95% CI

|  |  | Neurological diseases |  |  |  | Other diseases for reference |  |  |
| --- | --- | --- | --- | --- | --- | --- | --- | --- |
|  |  | Alzheimer's disease | Epilepsy | Parkinson's disease | Migraine | Hypertension | Diabetes | Dyslipidemia |
| Covariates | Age (in August 2020) | 1.001<br>( 0.997~1.005 ) | 1.001<br>( 0.997~1.005 ) | 1.001<br>( 0.997~1.005 ) | 1.001<br>( 0.997~1.005 ) | 0.999<br>( 0.994~1.003 ) | 1.001<br>( 0.997~1.005 ) | 0.999<br>( 0.995~1.003 ) |
|  | Sex (female) | 1.27<br>( 1.161~1.391 ) * | 1.271<br>( 1.161~1.392 ) * | 1.27<br>( 1.16~1.391 ) * | 1.269<br>( 1.159~1.389 ) * | 1.302<br>( 1.189~1.427 ) * | 1.25<br>( 1.141~1.369 ) * | 1.26<br>( 1.151~1.38 ) * |
|  | Being a visit for the disease of interest | 0.885<br>( 0.499~1.569 ) | 1.044<br>( 0.876~1.244 ) | 0.853<br>( 0.541~1.346 ) | 1.632<br>( 0.847~3.144 ) | 1.364<br>( 1.228~1.516 ) * | 0.662<br>( 0.555~0.789 ) * | 1.343<br>( 1.204~1.498 ) * |

Abbreviations: SHI, society-managed, employment-based health insurance association; NHI, National Health
Insurance; CI, confidence interval; AD, Alzheimer's disease; PD, Parkinson's disease; HTN, hypertension; DM,
diabetes; DL, dyslipidemia

## 4. Discussion

In this study, we analyzed an administrative claims database to investigate the influence of the COVID-19 pandemic on ambulatory care for chronic neurological diseases in Japan in early 2020. The key findings are as follows: (1) regular visits for epilepsy and PD mildly decreased since April 2020 in NHI but not in SHI; (2) the frequency and proportion of telephone re-examination use in ambulatory care visits for neurological diseases generally varied according to facility; (3) the telephone re-examination utilization rate generally reached its peak in May 2020, given the lifting of the nationwide ban in March 2020, at approximately 5% of the visits at most; and (4) there was no specific increase in the utilization rate of telephone re-examination during regular visits for neurological diseases of interest. A major advantage of this study is that it used the DeSC database, which has higher accessibility and analyzability than the national database (NDB), to investigate recent claims data, despite its weakness of smaller sample size. Our results thus provide basic insights about the trends in ambulatory care for chronic neurological disease patients at outpatient clinics during the first half of 2020, setting the stage for further research on the influence of COVID-19 on patients with neurological diseases.

In line with an earlier study by the Japan Medical Association [4], we confirmed the decline in regular visits to outpatient clinics for the sake of medicine prescriptions for any disease (Figure 2A, Table 3), both in SHI and NHI, presumably reflecting one of the social responses to the COVID-19 pandemic—refraining from stepping out to reduce the risk of COVID-19 infection. For patients who need regular visits to outpatient clinics for their chronic diseases given their requirement of continuous medications, increasing the number of prescription days would contribute to the decrease in the monthly visit count. Meanwhile, the use of telephone re-examination may have led to an increase in the monthly visit count because many of the facilities available for regular visits by telephone re-examination self-regulated the prescription days, making it shorter than that in the case of in-person visits, for example, 30 days or so (authors’ observation). Considering the low utilization rate of telephone re-examination (5% or less at most: Figure 4), increasing the prescription days may have been one of the causes of the decrease in regular outpatient clinic visits.

A decline in the monthly regular visits for epilepsy or PD since April 2020 was observed only in NHI but not in SHI (Table 3), which would be mainly due to the difference between the two groups in age distribution, that is, the baseline number/proportion of patients with epilepsy or PD was lower in the younger SHI population than in the older NHI population (Table 1). Further, we suspect some facility-dependent reasons as well, because patients with neurological diseases in general would more likely seek care from neurologists or at facilities specialized for the treatment of neurological diseases. For example, the “Top-1” facility in Figure 3C seemed to have received many visits by epilepsy patients before the COVID-19 pandemic, temporary shutting down in April 2020 possibly due to nosocomial outbreak or any other reason, and subsequently restarting its outpatient clinic along with telephone re-examination. Such a temporary closure of outpatient clinics, as a result of the variation caused by chance during the COVID-19 pandemic, may have contributed to the decrease in visits to NHI but not in SHI.

Until March 2020, the use of telephone re-examination in outpatient clinics in Japan had been largely limited, even for regular visits for chronic diseases. On February 28, 2020, the MHLW announced that they are lifting the nationwide ban on telephone re-examination for outpatient clinics as one of the exceptional measures against the COVID-19 pandemic [5]. Aside from the initial plans to prevent COVID-19 infection risks, the consequences of lifting the ban on telephone re-examination, in terms of actual efficacy and safety, has hardly been validated in Japanese clinical settings. Although currently, there are no established guidelines, careful evaluation will be needed beforehand to determine the diseases or cases for which the use of telephone re-examination may be especially inappropriate [10, 24]. For example, patients with epilepsy and poor sleep quality were found to have an increased risk of worsening seizures during COVID-19 in Italy [24]. Patients with neurological diseases, such as neurodegenerative diseases, often require in-person neurological examinations at outpatient clinics to be evaluated for disease progression or their current disease status, so that telephone re-examination in lieu of ambulatory care may be less appropriate for PD [10] compared to other neurological diseases such as epilepsy or migraine.

As with the case of decrease in outpatient visits, the reasons for using telephone re-examination are not confined to patient-specific factors. In light of the nosocomial outbreak of COVID-19 [23], some clinics have been forced to temporarily switch to telephone re-examination completely. The Top-1 facility in Figure 3C may be considered as an example, since the outpatient visits in this facility temporarily decreased to 0 in April 2020. A high variance (represented by CV) across different facilities was observed in the rate of telephone re-examination for epilepsy (Figure 3C), explaining the high rate of telephone re-examination used by epilepsy patients since May 2020 in NHI (Figure 4D). When subtracting the facility-dependent variance, the utilization rate of telephone re-examination for epilepsy in NHI was not very high (Figure 4E), which indicates that telephone re-examination was used in approximately 5% or less of visits for chronic neurological diseases.

Overall, combined with other results that the mild decrease in outpatient visits for ambulatory care for chronic neurological diseases, especially re-examination for neurological diseases, was not associated with the use of telephone (Table 4), our study showed that the impact COVID-19 pandemic had on the ambulatory care for some chronic neurological disease had been relatively limited during the first half of 2020 among the insurers examined in this study, in terms of the frequency or type of outpatient visits. The generalizability of the obtained results is limited, given the relatively smaller size of claims database by nature; however, the degree of decline in patient visits or the utilization rate of telephone re-examination was similar to that of an earlier questionnaire survey [4], thereby supporting a certain level of validity in the results obtained in the current study.

Our study has several limitations. First, the disease definition of neurological diseases used in this study has not always been validated, and it is impossible to return to the original electronic medical record for validation. The disease definitions used in this study are focused more on the content of prescriptions, which could lead to some underestimation of the actual diseases of interest. For example, patients with AD dementia do not always take anti-cholinesterase drugs because of their positive side effects or negative main effects. PD patients in their earliest stage would not be detected by our definition because they often do not receive any anti-PD medications. Furthermore, there is another concern about the overestimation of neurological diseases, especially in the case of epilepsy, since anti-epileptic drugs can sometimes be used for other indications. For instance, valproic acid is often used for bipolar disorder or migraine prophylaxis, and carbamazepine can also be prescribed for bipolar disorder or trigeminal neuralgia. Clonazepam is another anti-epileptic drug that is frequently used to treat symptoms of movement disorders.

Moreover, because the DeSC database used in this study does not disclose the details of the regions and localities of the insurers, NHI and SHI, we were unable to account for regional differences caused by differences in the timing and the extent to which the COVID-19 pandemic influenced the local medical systems across Japan, which may be one of the important confounders. In addition, the insurers in this database do not cover late-stage older adults aged 75 years or older, which means many neurological diseases, where the age of onset is typically late, cannot be examined sufficiently. These database specifications limit the generalizability of the current results; therefore, further validation studies using nationwide claims data (e.g., NDB) will be necessary to obtain a more robust conclusion. Lastly, because this study focuses on examining the outpatient clinic by telephone, the analysis was conducted only for claims reimbursed to medical institutions with less than 200 beds; therefore, our study did not take into account the differences in patient or disease demographics across medical institutions of different scales.

For future research, there are several additional questions that could not be addressed in this study. For example, we did not assess the first outpatient visit, the decline in which could also lead to a decrease in the subsequent outpatient visits in the long term. This is because we could not identify the first outpatient visit adequately due to our medication-based disease definition; we consider that many patients with neurological diseases may not receive prescription during their first visit. In addition, the cases where the patient’s family member visits on behalf of the patient could not be distinguished by the database as it is simply claimed as an in-person visit fee, although virtually this is an alternative to in-person re-examination and is similar to telephone re-examination because doctors do not examine the patients’ conditions directly. Furthermore, telemedicine need not always be confined to the use of telephone, but could also include video-visits using different electronic devices (e.g., smartphones, tablets, PCs, etc.) [8]. These are not distinguishable in the claims data, and thus, the difference between these devices or modalities should also be considered.

## 5. Conclusions

In conclusion, our study suggests that, in Japan, the impact of the COVID-19 pandemic on ambulatory care for certain chronic neurological diseases was relatively limited during the first half of 2020 in terms of the frequency or type of outpatient visits.

## Data Availability

The data used in this study was obtained from DeSC database by application to DeSC Healthcare, Inc.

## Acknowledgement

We are grateful to DeSC Healthcare, Inc. and their staff for kindly providing us with the database for this analysis.

## Figure legends

Supplemental Figure 1. Serial increase in the monthly rate (%) of unique patients who underwent telephone re-examination among all the unique patients with re-examination visits at the outpatient clinic Serial changes between January and July 2020. The vertical dotted line represents March 2020. Please note that the eligible period is until June 2020 in SHI (A-C) and until July 2020 in NHI (D-F). In addition, the y-axis scale is 0-20 only in (D), whereas it is 0-5 in others.

Abbreviations: SHI, society-managed, employment-based health insurance association; NHI, National Health Insurance; AD, Alzheimer’s disease; PD, Parkinson’s disease; HTN, hypertension; DM, diabetes; DL, dyslipidemia

