## Supplementary figures and images for "The impact of COVID-19 pandemic on the provision of ambulatory care for patients with chronic neurological diseases in Japan: evaluation of an administrative claims database"

### Supplemental Figure 1

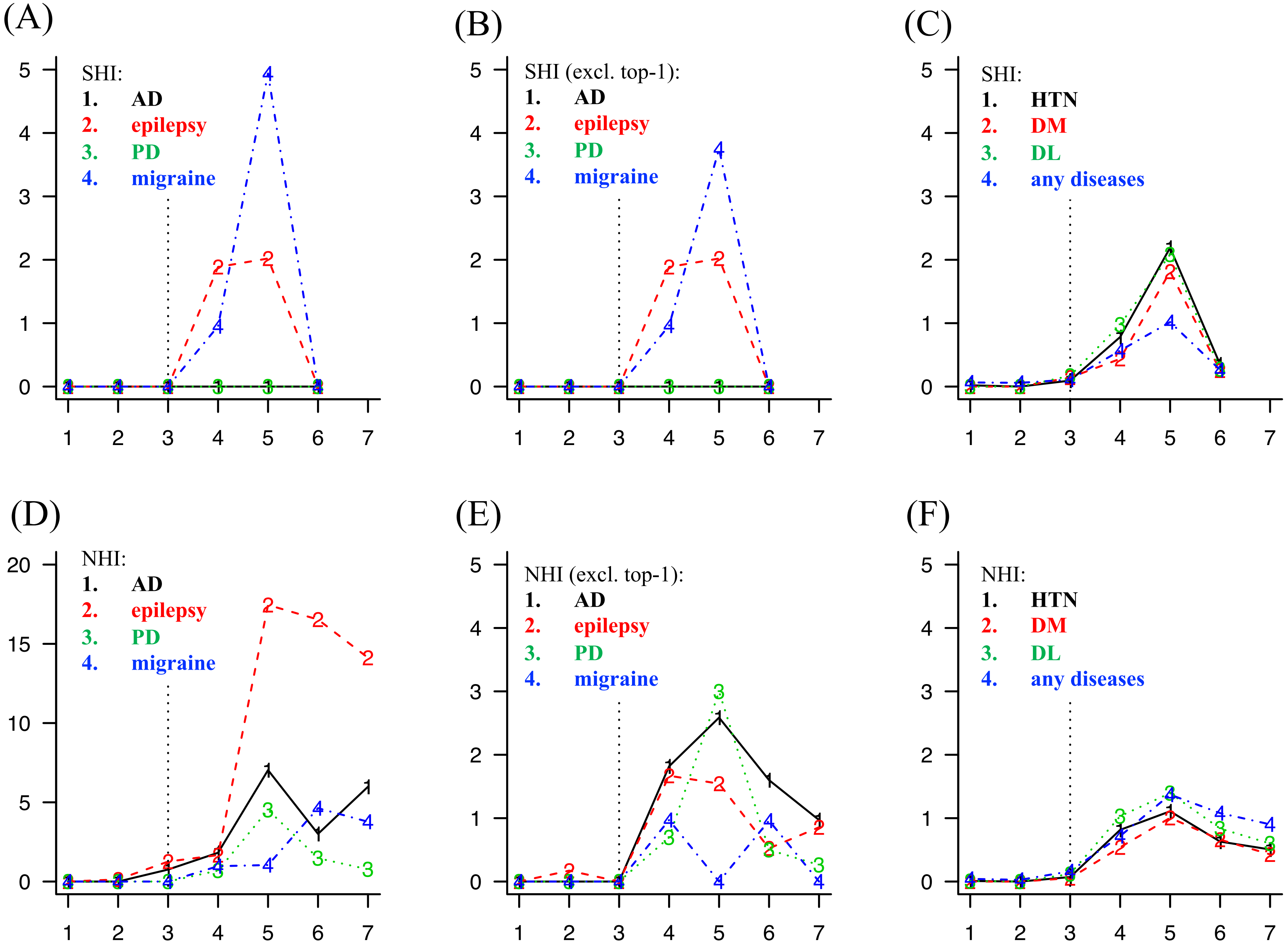
