## Supplemental Table 1 for "The impact of COVID-19 pandemic on the provision of ambulatory care for patients with chronic neurological diseases in Japan: evaluation of an administrative claims database"

Supplemental Table 1. Decrease in the number of unique patients who visited the outpatient clinic since April 2020: RR and ITSA

(A). RR results and 95% CI

| (by unique patients) | SHI | NHI |
| --- | --- | --- |
| Disease | Minimum RR | Minimum RR |
| Alzheimer's disease | 0.476 ( 0.133 ~ 1.706 ) , May | 0.904 ( 0.749 ~ 1.091 ) , April |
| Epilepsy | 1.009 ( 0.825 ~ 1.235 ) , June | 0.877 ( 0.811 ~ 0.947 ) * , April |
| Parkinson's disease | 0.85 ( 0.541 ~ 1.335 ) , June | 0.891 ( 0.809 ~ 0.981 ) * , May |
| Migraine | 1.067 ( 0.893 ~ 1.274 ) , March | 0.985 ( 0.822 ~ 1.18 ) , April |
| Hypertension | 0.931 ( 0.905 ~ 0.958 ) * , June | 0.967 ( 0.954 ~ 0.98 ) * , May |
| Diabetes | 0.911 ( 0.862 ~ 0.963 ) * , June | 0.964 ( 0.938 ~ 0.99 ) * , May |
| Dyslipidemia | 0.934 ( 0.905 ~ 0.965 ) * , June | 0.96 ( 0.945 ~ 0.976 ) * , May |
| Any diseases | 0.866 ( 0.856 ~ 0.876 ) * , April | 0.934 ( 0.927 ~ 0.942 ) * , May |

(B). Results of ITSA as an event intercept and its 95% CI

| (by unique patients) | SHI | NHI |
| --- | --- | --- |
| Disease | Impact in April 2020 | Impact in April 2020 |
| Alzheimer's disease | 0.668 ( 0.181 ~ 2.319 ) | 0.928 ( 0.781 ~ 1.102 ) |
| Epilepsy | 1.088 ( 0.905 ~ 1.308 ) | 0.9 ( 0.84 ~ 0.964 ) * |
| Parkinson's disease | 0.744 ( 0.496 ~ 1.11 ) | 0.813 ( 0.744 ~ 0.888 ) * |
| Migraine | 1.021 ( 0.845 ~ 1.233 ) | 0.878 ( 0.734 ~ 1.05 ) |
| Hypertension | 0.901 ( 0.877 ~ 0.925 ) * | 0.968 ( 0.955 ~ 0.98 ) * |
| Diabetes | 0.885 ( 0.841 ~ 0.932 ) * | 0.957 ( 0.933 ~ 0.981 ) * |
| Dyslipidemia | 0.893 ( 0.867 ~ 0.92 ) * | 0.962 ( 0.947 ~ 0.976 ) * |
| Any diseases | 0.762 ( 0.754 ~ 0.771 ) * | 0.903 ( 0.896 ~ 0.91 ) * |

Abbreviations: RR, relative risk; ITSA, interrupted time-series analysis; CI, confidence interval; SHI, society-managed, employment-based health insurance association; NHI, National Health Insurance.
